# Understanding the Impact of Digital Technology on the Well-being of Older Immigrants and Refugees: A Scoping Review Protocol

**DOI:** 10.1101/2023.08.01.23293451

**Authors:** Prince Chiagozie Ekoh, Tochukwu Jonathan Okolie, Fidel Bethel Nnadi, Oluwagbemiga Oyinlola, Christine A. Walsh

## Abstract

Although, scholarly reports show that some older adults utilise digital technology to enhance their well-being, a significant number of older adults are digitally alienated. This is complicated for older immigrants and refugees, whose situations present a peculiar challenge, requiring digital technology for improved quality of life. Hence, this scoping review seeks to understand the impact of digital technology on the well-being of older immigrants and refugees. Arksey & O’Malley’s five-stage framework will guide the review. The following databases: Social Work Abstract, Social Service Abstracts, Abstracts of Social Gerontology, International Bibliography of the Social Sciences (IBSS), etc., will be searched. Citations from the databases will be exported to Zotero to eliminate duplicate articles. These citations will be subjected to two screening levels. For the first screening, citations will be exported from Zotero to Rayyan QCRI© for title and abstract review. After that, all the authors will conduct full-text reviews of all articles included. The Preferred Reporting Items for Systematic Reviews and Meta-Analyses extension for Scoping Reviews (PRISMA-ScR) will be utilised in describing and documenting the inclusion and exclusion process. This scoping review will engender an improved understanding of the implications of digital technology on the well-being of older immigrants.

## Introduction

The adoption and use of digital technology have transformed how people communicate, access information and engage in various activities. Older adults are among the population that can significantly benefit from using digital technology, with some studies indicating that it improves older adults’ quality of life (Francis et al., 2019; Gill-Clavel et al., 2022). The COVID-19 pandemic, which exposed older adults to health risks, loneliness, social isolation, and misinformation, heightened the need for the adoption of digital technology by older adults (Nimrod, 2020; Rorai & Perry, 2020; Xie et al., 2020). For older immigrants and refugees, the use of digital technology can play a significant role in their overall well-being by providing access to essential services, resources (Vie et al., 2020) and social connections (Gill-Clavel et al., 2022) which might not be available through the traditional means (Neves, 2013).

Evidence suggests that digital technology is essential for the survival (Khoir et al., 2014) and resettlement of immigrants and refugees (Goodall et al., 2020; Tinghög et al., 2010). The healthcare, employment, inclusion, housing, and education information needs of immigrants and refugees can be met through the internet and digital technology (Broadbent & Papadopoulos, 2013). Digital technology is now critical for economic, social and political participation, increasing social inclusion, community building and social capital development (Broadbent & Papadopoulos, 2013).

Irrespective of the growing importance of digital technology in our daily lives, with new ongoing advancements in artificial intelligence, as well as other virtual realities, etc., promising potential benefits for older adults (Francis et al., 2019; Gill-Clavel et al., 2022), they are also at risk of being digitally excluded (Adam et al., 2019; Ekoh et al., 2021; Xie et al., 2020). This is notably worse for older migrants who are distanced from their loved ones because of migration (Adam et al., 2019; Correa-Velez et al., 2013; Warnes & Williams, 2006; Tinghög, 2010), and thus should enjoy the connection opportunities provided by digital technology but find themselves at the wrong side of the ‘digital divide’. The intersection of age, language, level of education, social-economic status, culture, familiarity, and understanding of technology disproportionately affect older refugees’ adoption of digital technology, leading to digital exclusion (Bianco et al., 2010; Lin et al., 2020). Tinghög (2010) reported that in contrast to younger refugees who benefited from digital technology to connect with social networks, which decreased their depressive symptoms, older refugees in Sweden did not adopt digital technology to maintain social ties. Other scholars have shown how limited access to digital technology by older immigrants and refugees has limited their connection to their community, reduced social inclusion and limited their access to healthcare and community resources (Alam & Imram, 2015; Lin et al., 2020).

In addition to the denial of social bonds, which digital technology can facilitate with its psychosocial benefits, the lack of access to and utilisation of digital technology can put older immigrants and refugees at risk of exclusion from mainstream information, making their integration and access to resettlement distressing (Alam & Imram, 2015). For instance, older migrants in Western countries like Australia and Canada who experienced digital exclusion struggled with access to education, housing, employment and health (Alam & Imram, 2015; Caidi & Allard, 2005). Thus, understanding how literature has described the impact of digital technology on the overall well-being of older immigrants and refugees is warranted. Therefore, this scoping review aims to identify and map out existing literature on the impact of digital technology on the well-being of older immigrants and refugees. The scoping review will improve our understanding of how the adoption of digital technology has enhanced the living conditions of older immigrants and refugees. It will also expose areas needing more research focus as digital technology advances.

## Methods

The review will be guided by the five stages of Arksey & O’Malley’s (2005) framework, which includes:

### Identifying the research question(s)

The scoping review will be guided by the following research questions (1) what are the impacts of digital technology on the well-being of older immigrants and refugees? (2) How can digital technology be improved for the improved quality of life of older immigrants and refugees?

### Identifying relevant studies

Electronic databases of published peer review empirical studies will be searched to identify studies relevant to this review. The databases to be searched will include Social Work Abstract, Social Service Abstracts, Abstracts of Social Gerontology, International Bibliography of the Social Sciences (IBSS), Sociological Abstracts, Science Direct, Web of Science, PsycINFO, PubMed and Technology collection (ProQuest). The key terms that will be searched in the databases are in table 1 below. In collaboration with a librarian, the authors will determine these search terms. Grey literature will be excluded, given the difficulties in evaluating the findings for the purpose of the scoping review.

**Table 1.** Key terms and MeSH terms.

| Concept | Key or MeSH terms |
| --- | --- |
| Digital technology | “Digital technology” OR “Internet” OR “Software” OR “ICT” |
| Older adults | “Older adults” OR “Older people” OR “Older persons” OR “Seniors” OR “ag*ing” OR (MH “Aging”) OR (MH “Aged”) OR “elder*” OR “elderly |
| Immigrants | “Immigrants” OR “Refugees” OR “Older immigrants” OR “Older refugees” OR “Displaced older adults” OR “Displaced persons” |
| Wellbeing | “Wellbeing” OR “Quality of life” OR “Welfare” OR “Successful ageing” |

For all the database searches, the symbol \* will be used to allow the inclusion of varied word endings.

The criteria for inclusion comprise 1) Empirical studies containing the terms with the title, abstract and keywords, 2) Availability of full-text articles electronically, 3) Publication is in English, 4) Research focus on the impact of digital technology on the well-being of older immigrants and/or refugees, 5) Participants are older immigrants or refugee and 6) The age of the study population are 50 years and above. While the exclusion criteria include: 1) Gray literature, 2) Non-human studies, (3) Participants are younger (less than 60 years), (4) randomised control-trial, (5) epidemiology studies. When there is a misunderstanding about the inclusion criteria fitting into a study, the study author(s) will be contacted three times requesting clarifications and articles whose author(s) do not respond on the third attempt will be excluded.

### Selecting studies

When the data search is completed, all the citations will be exported to Zotero, a reference management software to eliminate duplicates. After eliminating the duplicates with Zotero, the citations will be exported to Rayyan QCRI© for the first screening level-title and abstract review. The abstract and title review will be done based on the abovementioned inclusion and exclusion criteria. In the second screening level, the authors will also use the inclusion and exclusion criteria to review the full texts of all articles included after the “title and abstract” review. The snowballing technique will be adopted to search the included articles’ references to ensure that no article is missed. This will continue until no new articles meeting the inclusion/exclusion criteria are identified. This process will be documented and presented using a flow chart.

### Data collection and charting

The article number, authors’ names, the title of articles, the journal including volume, issue and page number, year of publication, study countries or regions, countries of older migrants, older migrants host countries, sample size and gender distribution, study aim, hypothesis/ research question, type of study, (qualitative, quantitative, or mixed-method), research design, significant findings, recommendations, limitations, and future research suggestions will be extracted using Microsoft Excel Data-charting form.

### Summarising, synthesising and reporting the results

The selected articles’ metadata will be summarised, and the results will be reported using narratives, visuals (e.g., maps or diagrams), tables or a combination of the two methods. Following Arksey and O’Malley’s (2005) recommendation, the reporting process will be documented to avoid potential bias. We will adopt the Preferred Reporting Items for Systematic Reviews and Meta-Analyses extension for Scoping Reviews (PRISMA-ScR) developed to guide reporting of scoping reviews (Tricco et al., 2018) in describing and documenting the inclusion and exclusion process.

## Discussion

Digital technology has become integral to our daily lives, from communication to commerce, education, and entertainment. Diverse populations of older adults have benefited from the use of digital technology. It reduced social isolation and loneliness amongst institutionalised older adults during the height of the COVID-19 pandemic-induced social distancing (Rorai& Perry, 2020), connected older adults to community resources (Xie et al., 2020) and improved access to healthcare information (Goodall et al., 2014). However, due to digital exclusion, older immigrants and refugees may not take full advantage of the potential of digital technology due to language barriers, cultural differences, and digital literacy. Thus, this scoping review is considered a first step towards mapping out studies that will aid our understanding of the impact of digital technology on the well-being of older immigrants and refugees. The review can inform interventions and policies to address the digital exclusion of older immigrants and refugees, which will improve their well-being and be a potential tool for older immigrants and refugees to connect with their communities, access resources and services, and engage in a range of activities.

## Data Availability

All data produced in the present study are available upon reasonable request to the authors

## Notes

### Competing Interest Statement

The authors have declared no competing interest.

### Funding Statement

Nil

